# The Association Between Biomarkers and Clinical Outcomes in Novel Coronavirus (COVID-19) Pneumonia in a U.S. Cohort

**DOI:** 10.1101/2020.05.27.20115105

**Authors:** Shant Ayanian, Juan Reyes, Lei Lynn, Karolyn Teufel

## Abstract

**Background:** The global pandemic caused by COVID-19 remains poorly understood by clinicians. Identifying biologic markers associated with prognosis can help clinicians recognize disease severity.

**Objective:** To describe the association between D-dimer, CRP, IL-6, ferritin, LDH, and clinical outcomes in a cohort of COVID-19 patients treated on the inpatient medical service at a university hospital in Washington, DC.

**Design:** In this retrospective study, we included all adults admitted to the inpatient medicine service at George Washington University Hospital between March 12, 2020 and May 9, 2020 with laboratory confirmed COVID-19. Clinical and laboratory data were extracted from electronic medical records and compared between survivors not requiring ICU transfer, survivors requiring ICU transfer, survivors requiring intubation, and non-survivors.

**Key Results:** 299 patients were included in our study, of whom 69 required transfer to the ICU, 39 required intubation, and 71 died. Threshold values for IL-6 (≥50 pg/mL), D-dimer (≥3 mcg/mL), ferritin (≥450 ng/mL), CRP (≥100 mg/L), and LDH (1,200 u/L) were found to be statistically significant and independently associated with higher odd of clinical deterioration and death. Hypertension, CVA and heart disease independently had an increased risk of all three outcomes, while CKD had only an increased risk of death. Patient co-morbidities had no effect on the different biomarkers’ significant association with poor patient clinical outcomes, except cancer.

**Conclusion:** Laboratory markers of inflammation and coagulopathy can help clinicians identify patients who are at high risk for clinical deterioration, independent of clinically significant medical comorbidities.

## Introduction

A viral pneumonia of unknown cause detected in Wuhan, China was first reported to the World Health Organization (WHO) Country Office in China on 31 December 2019.^1^ In the United States, the virus was first seen in January 2020, in a traveler who returned from China.^2^ By February 2020, WHO had given the virus a name: the novel coronavirus 2019, shorthand COVID-19. In early March 2020, shortly after the first cluster of cases was diagnosed in the Western United States,^3^ WHO declared a pandemic.

New York City was the first major metropolitan area on the US East coast to see large numbers of COVID-19 patients. By early March, the virus was confirmed in the District of Columbia.^4^ Shortly thereafter, the first cases were diagnosed at George Washington University Hospital (GWUH) in Washington, DC. In the following 2 months, 299 cases of confirmed COVID-19 were admitted to our hospitalist service.

Previous studies of COVID-19 have identified various biologic markers associated with poor prognosis. An association between mortality and both elevated inflammatory markers and coagulation functional indices was first reported in a retrospective cohort study of 138 patients with COVID-19 in Wuhan.^5^ In this study, higher D-dimer and IL-6 levels were seen in patients requiring ICU care, in patients with ARDS, and in non-survivors. Also published in March 2020, a second retrospective analysis of 191 patients found that elevated levels of LDH, ferritin, D-dimer, and IL-6 were associated with mortality.^6^ This group reported that their patients with D-dimer greater than 1mcg/L at the time of admission had an 18-fold increase in the odds of in-hospital death. In a subsequent retrospective analysis conducted at Tongji Hospital, Tang et al demonstrated that of 183 COVID-19 patients, ≥ 70% of the non-survivors had an elevated D-dimer on admission.^7^ In a commentary on Tang’s data, Canadian Dr. Lillicrap concluded, “The observations of Tang and colleagues provide early evidence that enhanced vigilance is required to identify the emergence of DIC in 2019-nCoV pneumonia patients.”^8^

Based on these and other reports in the literature,^9^ clinicians in the US have been measuring a variety of biologic markers in COVID-19 patients. The clinical impact of these biological markers on outcomes in the U.S. population remains unclear. At GWUH, during the first few weeks of the COVID-19 pandemic, hospitalists typically measured markers of inflammation and coagulopathy, but no standardized protocol for laboratory testing had been established. Our team sought to examine the association between these markers and clinical outcomes of our COVID-19 patients, with the goal of identifying which markers had clinical utility in assessing prognosis.

## Methods

All adult (≥18 years of age) patients diagnosed with COVID-19 and admitted to the medicine service at GWUH between March 12, 2020 and May 9, 2020 were included in the study. Clinical, laboratory and outcome data were extracted from electronic medical records using a standardized data collection form. Age, sex, BMI, comorbidities, and medications were recorded. In addition to routine admission laboratory studies, C-reactive protein (CRP), D-dimer, interleukin 6 (IL-6), ferritin, and lactate dehydrogenase (LDH) were recorded. Further, the maximum oxygen requirements prior to transfer to intensive care unit, transfer to the ICU, necessity for mechanical ventilation, and discharge status were noted.

### Statistical Analysis

The data analysis was conducted using statistical software R. Given the sample size and the need for reliable markers for disease severity, the continuous variables were transformed into categorical variables. For IL-6, ferritin, CRP, and LDH, the median of each group was used as the threshold [IL-6 ≥ 60 pg/mL (normal range 0.0–15.5 pg/mL), ferritin ≥ 450 ng/mL (normal range 20–450 ng/mL), and CRP ≥ 90 mg/L (normal range 0.0–9.0 mg/L), and LDH is 1200 u/L (normal 400–800 u/L). For D-dimer (normal range 0.20–0.28 mcg/mL), a level of 3 mcg/mL was used. This level had been reported in the recent literature from China as having a higher specificity for predicting thrombi.^10^

After the key characteristics of the variables were studied, a logistic regression model was fitted with D-dimer, IL-6, LDH, ferritin, and CRP with ICU admission, intubation, and death sequentially. As all the coefficients of these markers were found to be statistically significant, the odds ratio of all three outcomes were calculated for patients with the categorical variables of IL-6, D-dimer, ferritin, LDH and CRP.

A stratification analysis was carried out with every one of the comorbid conditions recorded. These conditions were classified by the Elixhauser disease severity index and grouped into 8 categories of interest. A weighted AHRQ disease severity index was also calculated and later transformed into a binary variable with 1 indicating patients with a score higher than 10. Furthermore, a stratification with regards to sex and BMI>30 was carried out. The mean age of patients within each outcome category was also compared.

This study was approved by the Institutional Review Board at George Washington University.

### Results

A total of 299 patients with confirmed COVID-19 positive results were admitted to the hospitalist service at GWUH between March 12^th^ and May 9^th^, 2020. Of these patients, 200 had all the biomarkers of interest measured by their clinicians. The median follow-up period was 8 days. Sixty-nine patients (23%) were transferred to the ICU, thirty-nine patients (13%) required intubation and seventy-one patients (23.7%) died. The key laboratory findings for the 5 biomarkers can be found in table 1, and the comorbid conditions are illustrated in table 2. If any of the markers were measured a few times during the hospitalization, the peak value was used during analysis.

**Table 1:** Biomarkers studied with key characteristics

| Variable Studied | Median | Mean | Number of available results |
| --- | --- | --- | --- |
| IL-6 (normal range 0.0-15.5 pg/mL)<sup>i</sup> | 65 pg/mL | 206 pg/mL | 222 |
| D-dimer (normal range 0.20-0.28 mcg/mL) | 2.1 ng/mL | 4.1 mcg/mL | 248 |
| Ferritin (normal range 20-450 ng/mL) | 446 ng/mL | 880.9 ng/mL | 252 |
| CRP (normal range 0.0-9.0 mg/L) | 90 mg/L | 159 mg/L | 241 |
| LDH (normal range 600-800 units/L) | 974 units/L | 1250 units/L | 254 |

IL-6 = interleukin 6; CRP = C-reactive protein; LDH = lactate dehydrogenase
<sup>i</sup>IL-6 lab performed at: BN LabCorp, 1447 York Court Burlington, NC 27215; all other labs performed at George Washington University Hospital

**Table 2:** Comorbidities and outcomes, Odds Ratios

| Characteristics | Prevalence | ICU admission | Intubation | Death |
| --- | --- | --- | --- | --- |
| Sex = M | 54% | 2.6 (1.4, 4.6)* | 2.4 (1.1, 5.0)* | 1.4 (1.0, 2.2) |
| BMI >30 | 29% | 1.2 | 1.3 | 0.6 |
| HTN | 68% | 2.4 (1.2, 4.5)* | 2.8 (1.1, 7.0)* | 2.8 (1.4, 5.5)* |
| Diabetes | 46% | 1.3 | 1.7 | 1.2 |
| Cardiac disease | 46% | 1.9 (1.8, 3.3)* | 2.0 (1.0, 4.2)* | 3.6 (2.0, 6.3)* |
| CVA | 17% | 1.9 (1.3, 3.0)* | 3.2 (1.6, 6.4)* | 4.5(2.5, 8.0)* |
| COPD | 29% | 1.2 | 1.3 | 1.7 |
| CKD | 29% | 1.5 | 0.9 | 2.3 (1.4, 4.0)* |
| HIV | 4% | NA | NA | 0.27 |
| Cancer | 6% | 1.6 | 0.9 | 1.5 |

\*Indicates significance at p value of 0.05

Univariate logistic regression models yielded statistically significant estimates for IL-6 ≥ 60 pg/mL, D-dimer ≥ 3 mcg/mL, CRP ≥ 90 mg/L, ferritin ≥ 450 ng/mL and LDH ≥ 1200 u/L for ICU admission and intubation, and death. The odds ratio of the three outcomes with the different markers is illustrated in table 3. HTN, heart disease, and neurologic disorders had significant association with ICU admission, intubation and death. The other medical conditions were not significant, except for CKD having a significant association with death. Male sex had a higher odds ratio of intubation and ICU transfer than female sex. A separate stratified analysis was conducted for every marker with respect to the comorbid conditions to eliminate any confounding effect. The Mantel-Haenszel test for heterogeneity was not significant except for cancer (hematologic and solid malignancies) with respect to D-dimer, and sex with respect to IL-6. The adjusted OR are thus reported in table 3.

**Table 3:** Odds Ratios for all Biomarkers with 95% CI.

| **Biomarker** | **OR for ICU admission** | **OR for Intubation** | **OR for Death** |
| --- | --- | --- | --- |
| IL-6 ≥ 50 pg/mL (0.0-15.5 pg/mL) | 5.9 (2.7, 13.1)* | 4.6 (1.7, 12.4)* | 5.4 (2.4, 11.8)* |
| D-dimer ≥ 3 mcg/mL (0.20-0.28 mcg/mL) | 7.3 (3.8, 14.0)* | 8.2 (3.4, 19.8)* | 7.5 (3.8, 14.6)* |
| Ferritin ≥ 450 ng/mL (20-450 ng/mL) | 6.8 (3.4, 13.7)* | 4.0 (1.8, 8.8)* | 5.1 (2.6, 10.0)* |
| CRP ≥ 100 mg/L (0.0-9.0 mg/L) | 8.6 (4.1, 18.1)* | 6.5 (2.6, 16.3)* | 6.0 (3.0, 12.0)* |
| LDH ≥ 1200 (600-800 units/L) | 4.5 (2.5, 8.1)* | 5.4 (2.5, 11.8)* | 8.0 (4.1, 15.3)* |

IL-6 = interleukin 6; CRP = C-reactive protein; LDH = lactate dehydrogenase; OR = odds ratio
\*Indicates a p value of less than 0.05 (Fisher's exact test)

## Discussion

Although population-based studies have identified risk factors for poor prognosis in COVID-19, the clinical course of individual patients infected with the virus is highly variable. Risk stratification of medical comorbidities as well as biologic markers predicting clinical deterioration are needed for the U.S. Population.

In our cohort of patients, the only comorbidities associated with poor outcomes across all three endpoints were history of hypertension, CVA, and heart disease. Chronic kidney disease was associated with an increased odds of death. Other analyses have demonstrated an association between obesity and diabetes as having poor outcomes in hospitalized patients with COVID-19 infection.^5,6,11^ We suspect that our cohort of patients was too small to detect any other significant comorbidities.

Our retrospective study did identify an association between biomarkers and clinical deterioration as well as death. Elevated levels of IL-6, D-dimer, CRP, LDH, and ferritin all had an independent increased risk for the clinical outcomes assessed (ICU admission, invasive ventilatory support and death), which were statistically significant. Specifically, in our cohort, these biomarkers remained statistically significant in patients with hypertension and cardiovascular disease. These associations were also independent of any patient co-morbidities, except for cancer which appeared to have an effect on D-dimer. However, the prevalence of cancer in this population was low. Thus, a higher number of patients with this condition is required to confirm the confounding effect of cancer on D-dimer.

Of further significance is that our cohort of COVID-19 patients admitted to the hospitalist service revealed that the highest odds of death occurred when the LDH level was greater than 1200 u/L and a D-dimer level greater than 3 mcg/mL. Though more modest, all other biomarkers including IL-6, ferritin, and CRP also carried a statistically significant increased odds of death. These findings appear to corroborate that the inflammatory indices of IL-6, CRP, and ferritin are independently associated with patients becoming critically ill, requiring ventilatory support, and dying. It remains unclear whether these inflammatory indicators are biologic markers of disease or mediators of the hypothesized ‘cytokine storm,’ in which hyperinflammation and multi-organ disease arise through excessive cytokine release from uncontrolled immune activation.^12^ However, there are no studies to aid in contextualizing the significance of inflammatory biomarkers. It is possible these markers predict clinical course; if so, they could inform therapeutic interventions rather than simply demonstrate a consequence of the disease. Further study on the timing and frequency of measuring inflammatory biomarkers, and the value of trending them as it relates to clinical outcomes, is clearly warranted.

In our cohort, the odds of intubation were the highest with a D-dimer greater than 3 mcg/ml. These patients had a higher likelihood of clinically deteriorating and transferring to the ICU, requiring mechanical ventilatory support, and greater odds of death. Our decision to use a D-dimer threshold of 3 mcg/mL, rather than the median of 2.1 mcg/mL, was based on a study pointing to a D-dimer threshold of 3 mcg/mL as having a higher specificity for identifying patient with thrombotic complication.^10^

The clinical predictive value of an elevated D-dimer corroborates findings observed in prior studies (Zhou’s et, al; Tang et al; Wu et. Al) where an elevated D-dimer was associated with an increased risk of needing ventilator support and in-hospital mortality. Limited histopathologic and autopsy data show evidence of exudative inflammation, small vessel occlusion and a thrombotic microangiopathy suggestive of a hypercoagulable state.^13,14,15^ In addition, a single retrospective study has demonstrated mortality benefit in patients with high D-dimer levels (6 times the upper limit of normal) who were treated with prophylactic doses of LMWH.^16^ Of note, the mortality rate in their patients was substantially higher than that observed in our cohort, suggesting that we may be already achieving a benefit from the DVT prophylaxis that is routinely used in the US, in contrast to the clinical practice in China. The cohort of patients that Tang et al reviewed were not routinely treated with chemoprophylaxis for prevention of DVT. Whether therapeutic anticoagulation with heparin would yield greater benefit has yet to be demonstrated and deserves further clinical study.

Our study has some limitations. First, due to the retrospective study design, not all laboratory tests were done on all patients and, as a result, the role of an individual marker might be overestimated. Selection bias could overestimate the utility of these biomarkers. Our cohort did not include COVID-19 patients who were evaluated in the emergency room and deemed to be clinically well enough to return home. If biomarkers in these discharged patients were also elevated, their predictive value would be diminished. However, checking inflammatory biomarkers is not the standard of care in the emergency department. Perhaps the clinical context (referred for hospitalization) enhances the benefit of these biomarkers for an inpatient team.

Despite these limitations, we believe these markers can aid clinicians in identifying hospitalized COVID-19 patients at risk of clinical deterioration. Our findings suggest that regularly checking IL-6, D-dimer, CRP, LDH, and ferritin has clinical utility in this respect, especially when these markers are above the cut-off values mentioned. Following these biomarkers could ensure closer monitoring of these patients and provide guidance and standardization in the allocation of increasingly scarce resources.

## Conclusions

Laboratory markers of inflammation and coagulopathy can help clinicians identify patients who are at high risk for clinical deterioration. Future directions of research could focus on the temporal variability of these biomarkers and outcomes in COVID-19 patients, as well as, the effect of treatment with full dose anticoagulation.

## Data Availability

Relevant de-identified data may be made available on a case by case basis

## Acknowledgements

Thank you to Dr. Linda Lesky for her valuable input and the R project for Statistical Computing (https://www.r-project.org/).

## References

1. The World Health Organization: Emergencies/Diseases/Coronavirus disease 2019/Events as they happen. https://www.who.int/emergencies/diseases/novel-coronavirus-2019/events-as-they-happen. Last accessed April 20, 2020.

2. McNamara, A. CDC Confirms First Case of Coronavirus in United States. CBS News. Published January 21, 2020. https://www.cbsnews.com/news/coronavirus-centers-for-disease-controlfirst-case-united-states/. Last accessed April 20, 2020.

3. Selsky, Andrew. Washington governor declares state of emergency over virus. AP News. Published Feb 29, 2020. https://apnews.com/f175d89567a26d59cab27725c9e8a0d7. Last accessed April 20, 2020.

4. Custis, A. A timeline of the DC region’s COVID pandemic. D.C. Policy Center. Published March 24, 2020. https://www.dcpolicycenter.org/publications/covid-19-timeline/. Last accessed April 20, 2020.

5. Wang D, Hu B, Hu C, et al. Clinical characteristics of 138 hospitalized patients with 2019 Novel Coronavirus Infected Pneumonia in Wuhan, China. JAMA. Feb 2020;323(11):1061–1069.

6. Zhou F, Yu T, Du R, et al. Clinical course and risk factors for mortality of adult inpatients with COVID-19 in Wuhan, China: a retrospective cohort study. Lancet. March 2020;395(10229):1054–1062.

7. Ning T, Dengju L, Xiong W, Ziyong S. Abnormal Coagulation Parameters Are Associated with Poor Prognosis in Patients With Novel Coronavirus Pneumonia. J Thromb Haemost. April 2020;18(4):844–847.

8. Lillicrap, D. Disseminated Intravascular Coagulation in Patients With 2019-nCoV Pneumonia. J Thromb Haemost. 2020 Apr;18(4):786–787.

9. Min, C., Cheon, S., Ha, N. et al. Comparative and kinetic analysis of viral shedding and immunological responses in MERS patients representing a broad spectrum of disease severity. Sci Rep. May 6, 2016.

10. Cui, S., Chen, S., Li, X., Liu, S. and Wang, F. Prevalence of venous thromboembolism in patients with severe novel coronavirus pneumonia. J Thromb Haemost. 2020. Accepted Author Manuscript.

11. Richardson S, Hirsch JS, Narasimhan M, et al. Presenting Characteristics, Comorbidities, and Outcomes Among 5700 Patients Hospitalized With COVID-19 in the New York City Area. JAMA. Published online April 22, 2020. doi:10.1001/jama.2020.6775

12. Henderson, L., Canna, S., Shulert, Grant., Volpi, S., Lee, et. al. On the alert for cytokine storm: Immunopathology in COVID-19, Arthritis & Rheumatology. April 15, 2020.

13. Fox, S., Akmatbekov, A., Harbert, J., et. al. Pulmonary and Cardiac Pathology in Covid-19: The First Autopsy Series from New Orleans. BMJ. April 10, 2020.

14. Yao XH, Li TY, He ZC, et al. A pathological report of three COVID-19 cases by minimally invasive autopsies. Chinese Journal of Pathology. March 15, 2020; 49(0):E009.

15. Luo W, Yu H, Gou J, et al. Clinical pathology of critical patient with novel coronavirus pneumonia (COVID-19) Pre-print. March 9,2020.

16. Ning Tang, Huan Bai, Xing Chen, Jiale Gong, Dengju Li, Ziyong Sun. Anticoagulant treatment is associated with decreased mortality in severe coronavirus disease 2019 patients with coagulopathy, Journal of Thrombosis and Haemostasis. March 27, 2020.

